# Investigation of acute encephalitis syndrome with implementation of metagenomic next generation sequencing in Nepal

**DOI:** 10.1101/2023.03.21.23286552

**Authors:** R Shrestha, N Katuwal, D Tamrakar, CM Tato, M Vanaerschot, V Ahyong, J Gil, SK Madhup, BP Gupta, R Jha

**Author notes:** These authors contributed equally to this work (R Shrestha, N Katuwal).

## Abstract

**Background:** The causative agents of Acute Encephalitis Syndrome remain unknown in 68-75% of the cases. In Nepal, the cases are tested only for Japanese encephalitis, which constitutes only about 15% of the cases. However, there could be several organisms, including vaccine-preventable etiologies that cause acute encephalitis, when identified could direct public health efforts for prevention, including addressing gaps in vaccine coverage.

**Objectives:** This study employs metagenomic next-generation-sequencing in the exploration of infectious etiologies contributing to acute encephalitis syndrome in Nepal.

**Methods:** In this study, we investigated 90, Japanese-encephalitis-negative, banked cerebrospinal fluid samples that were collected as part of a national surveillance network in 2016 and 2017. Randomisation was done to include three age groups (<5-years; 5-14-years; >15-years). Only some metadata (age and gender) were available. The investigation was performed in two batches which included total nucleic-acid extraction, followed by individual library preparation (DNA and RNA) and sequencing on Illumina iSeq100. The genomic data were interpreted using Chan-Zuckerberg-ID and confirmed with polymerase-chain-reaction.

**Results:** Human-alphaherpesvirus-2 and Enterovirus-B were seen in two samples. These hits were confirmed by qPCR and semi-nested PCR respectively. Most of the other samples were marred by low abundance of pathogen, possible freeze-thaw cycles, lack of process controls and associated clinical metadata.

**Conclusion:** From this study, two documented causative agents were revealed through metagenomic next-generation-sequencing. Insufficiency of clinical metadata, process controls, low pathogen abundance and absence of standard procedures to collect and store samples in nucleic-acid protectants could have impeded the study and incorporated ambiguity while correlating the identified hits to infection. Therefore, there is need of standardized procedures for sample collection, inclusion of process controls and clinical metadata. Despite challenging conditions, this study highlights the usefulness of mNGS to investigate diseases with unknown etiologies and guide development of adequate clinical-management-algorithms and outbreak investigations in Nepal.

## Background

Acute Encephalitis Syndrome (AES) is defined by acute onset of fever and a change in mental status (including symptoms such as confusion, disorientation, coma, or inability to talk) and/or new onset of seizures (excluding simple febrile seizures) in a person of any age at any time of year.^1^ This term was coined by World Health Organization (WHO) in 2008.^1^ Globally, based on various studies, the incidence of AES has ranged from 3.5 to 7.4 per 100,000 patients-years, with a higher incidence among children.^2^

The patients suffering from AES usually present acute onset of fever and altered sensorium. This is followed by rapidly worsening clinical conditions and death.^3^ The survivors can suffer from long term health issues, including neurological sequelae.^3,4^ The etiologies behind AES can be grouped under infective and non-infective categories, with the infective category comprising of a broad range of organisms (bacteria, virus, parasites).^2,5^ The causative agents of AES also vary with season and geographic location.^6^ Research has shown that the etiologies of AES remain unknown in 68-75% of the cases, while Japanese encephalitis (JE) constitutes about 15% of the cases.^7–10^ The landscape of AES, in terms of etiology, has changed in India as well, where outbreak investigations and surveillance studies have increasingly reported non-JEV etiologies.^11^

In Nepal, JE is majorly associated with mortality and morbidity among children.^12^ Therefore, since 2004, the Ministry of Health and Population of Nepal, supported by the Office of Infection Prevention Division, World Health Organisation (WHO), has integrated JE surveillance with Acute Flaccid Paralysis, Neonatal Tetanus, and Measles in its National Surveillance Network.^13^ Until 2011, over 23,000 AES cases were reported by the surveillance network.^14^ Due to a lack of knowledge in etiology, AES cases are only tested for JE and clinical management is performed based on this result. The incidence of undiagnosed AES etiology contributes to a high rate of death and morbidity.^14^ There could be several etiologies, including vaccine preventable etiologies, that cause acute encephalitis, which upon identification could direct public health efforts for prevention, including expanded use of vaccines or addressing gaps in vaccine coverage. Herpes Simplex Virus (HSV), Varicella-Zoster Virus (VSV), Enterovirus, Adenovirus, and Rubella, as well as emerging pathogens such as Nipah, Chandipura and Chikungunya have all been reported as causative viral agents of AES, while Neisseria meningitidis, Streptococcus pneumoniae, Listeria sp, and Brucella have been reported as causative bacterial agents.^2,15,16, 17^

While molecular methods such as PCR require prior genetic information on causative agents, genomic methods such as metagenomic Next Generation Sequencing (mNGS) can simultaneously identify minute amounts of infections and co-infections of varying origin in a single investigation and assist in the investigation of transmission of such infections.^18, 19^ With the recent dramatic decrease in sequencing costs, this technology provides access to genomic information in a scale that can be implemented to fill gaps in routine clinical practice and address epidemiological questions. In addition to identification (identifying genotypes, virulence or pathogenesis), NGS provide information epidemiological investigation (comparative genomics, phylogenetic analysis).^20, 21, 22, 23^

This study, employing mNGS to explore the infective etiologies behind AES, complements a growing number of studies that have used a similar approach to investigate encephalitis, including in a LMIC context.^16,24,25,26^ The identification of such etiologies is an important step in developing effective prevention and treatment measures which in turn will reduce disability and morbidity.

## Methods

### Sample Collection and Selection

The investigation included a random selection of 90 retrospective cerebrospinal fluid (CSF) samples that were collected by WHO-IPD (World Health Organization-Immunization Preventable Diseases), throughout Nepal, as a part of the National AES Surveillance Network in collaboration with FWD (Family Welfare Department) in 2016 and 2017. These samples had been tested for JE at NPHL (National Public Health Laboratory) and stored at low but undocumented temperature. For this study, only those samples that tested negative for JE were selected. Randomisation was done to include three age groups of <5 years, 5-14 years, and >15 years. Only some metadata related to the subjects (age and gender) were known. Each sample was provided with unique study codes to maintain privacy.

### Nucleic Acid Extraction and mNGS

Total Nucleic Acid was extracted from the CSF samples using Zymo Quick-DNA/RNA™ Pathogen MiniPrep (R1042).

The total nucleic acid samples were aliquoted into two sub samples for RNA and DNA library preparation, respectively. The library preparations were done using NEB Library Prep Kit Ultra II RNA for RNA (New England Biolabs, E7770S) library preparation, and NEB Library Prep Kit Ultra II FS DNA for DNA library preparation (New England Biolabs, E7805S). The library preparation for the first 30 samples was done in a single batch (at Chan Zuckerberg Biohub, USA) while the remaining 60 samples were done in three batches of 20 samples each (at Dhulikhel Hospital Kathmandu University Hospital, Nepal). Negative extraction and library preparation controls were included in each batch. The library preparation included 10ng of input nucleic acid, followed by fragmentation, adapter ligation, cleanup (Solid Phase Reversible Immobilization beads), barcoding and amplification of library for 12-16 cycles. With subsequent library preparations, quality control was done using agarose gel electrophoresis and Tapestation 4200 platform from Agilent Technologies and later by qPCR using Kapa Illumina Library Amplification (KK2702) and Quantitation Complete Kit (KK4923). It was made sure that the length of DNA in the libraries was around 350-400bp and had concentration >1nM. In RNA library preparation, ERCC (External Control Controls Consortium, 4456740) RNA Spike-in controls were used as internal controls.

The libraries that passed quality control filters were pooled and run on an Illumina iSeq100 sequencer. The sequencing was performed for 2×146bp length using custom unique dual indices of 12 bp length. 5% PhiX was added as an internal control for sequencing. The loading concentration of pooled libraries was maintained at 100-120pM.

### Data Analysis

The analysis was performed on the CZ ID (formally known as IDSeq) platform developed by Chan Zuckerberg Initiative and CZ Biohub. CZID accepts raw sequencing data, perform host and quality filteration, followed by execution of assemblybased alignment pipeline.^27^The samples are analysed based on number of reads per million (Number of reads aligning to the taxon in the NCBI NR/NT database, per million reads sequenced), reads (Number of reads aligning to the taxon in the NCBI NT/NR database), contig number (Number of assembled contigs aligning to the taxon in the NCBI NT/NR database), id% and z-score. The samples were also visualized using a rpm heatmap where samples and controls are cross-matched against each other. Respective background models were created, from negative extraction and library preparation control, for RNA and DNA Libraries.

### PCR Confirmation

Human alphaherpes virus confirmation was done by qPCR (KAPA HiFi HotStart Ready Mix) using two primer sets: established primers (FP: 5’TGCAGTTTACGTATAACCACATACAGC 3’ and RP: 5’ AGCTGCGGGCCTCGTT 3’) and self-designed primers (FP: 5’ GACTCAAACACGTGCACCAC and RP: 5’ CCATCGCGTACAGCCTACAT 3’).^28^ The primer sets were designed using NCBI primer blast and Gene Script, then checked with Beacon Designer Free and Snap Gene Viewer.

Similarly, for confirmation of Enterovirus, modified protocol with established primers from Enterovirus Surveillance Guidelines were used to perform semi-nested Polymerase Chain Reaction (snPCR).^29^ The protocol followed visualisation of the bands, for confimration, in agarose (1.5%) gel electrophoresis.

## Results

### Subject Metadata

The samples selected for this study were banked, retrospective CSF samples collected in 2016 and 2017 with limited metadata such as age and gender. Out of the 90 subjects, 31 (34.4%) were female while the age distribution has been presented in table 1.

**Table 1:** Age Distribution between subjects with AES.

| SN | Age Group | Number |
| --- | --- | --- |
| 1 | <5 years | 23 |
| 2 | 5-14 years | 14 |
| 3 | >15 years | 53 |

The median age of the subjects infected with AES was 20 years (IQR: 4-79 years)

### Sample collection

The samples were collected in glass bottles without any preservative and transported to NPHL where they, first, had been tested for JE and subsequently stored at low temperature. As these samples were collected in 2016 and 2017 and banked, negative controls were not available during collection and transportation.

### Nucleic Acid Extraction and mNGS

The extracted nucleic acid had a concentration ranging from too low to detect to 222 ng/ul. As the analysis was done in two sets. Each set was processed for DNA and RNA Library Preparation and has been presented accordingly. Out of 90 samples, only two samples showed confirmed hits from Enterovirus B and Human alphaherpesvirus 2, respectively.

### mNGS of RNA Libraries

The results from RNA libraries showed some distinct organisms hit in CZID, and also provided a broad picture of the landscape of taxa across the samples. The following are heatmaps generated from through RNA library preparation.

In figures 1 and 2, we can see top hits of organisms in the heat map that shows various organisms which are seen at similar levels in the water controls as well. Nevertheless, Pseudomonas genus is seen in all of the samples including few negative controls. There was similar trend, in both sets, with other organism such as Sphingomonas, Acinetobacter, Escherichia and others.

**Figure 1:**
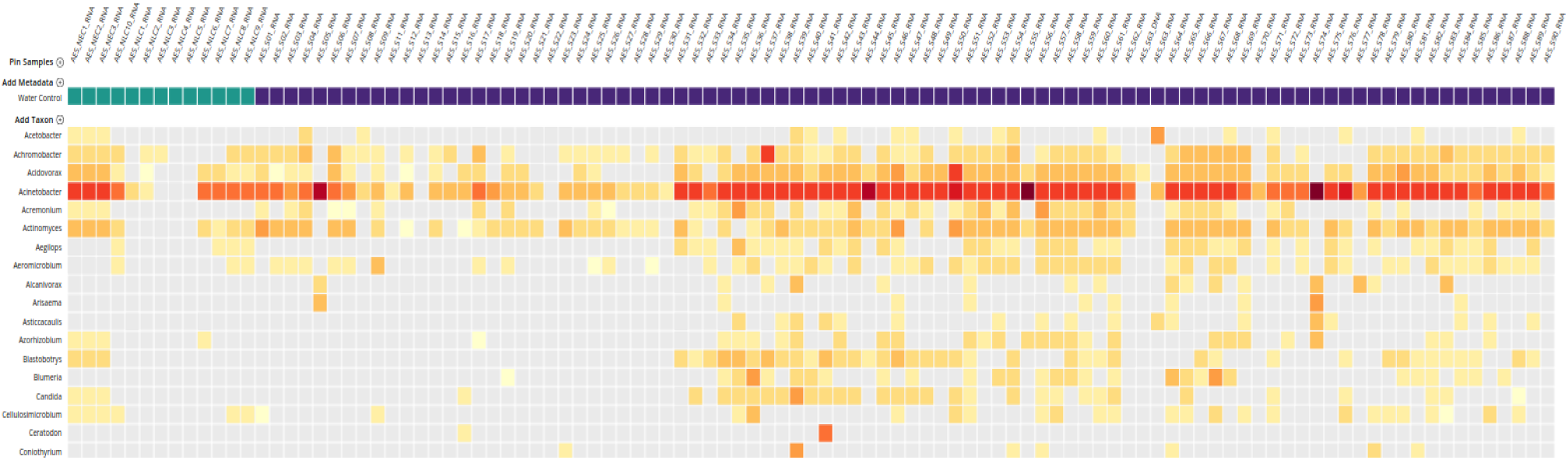
Heatmap depicting top hits from sequencing of RNA library for S01 to S90. The organisms (at the genus level) that were seen in the samples are shown on the left. The names of the samples are on the top of the heat map. The samples marked by green are water controls during extraction (NEC) and library preparation (NLC). The heatmap was generated using the threshold of NT rPM (nucleotide reads per million) >=10 and NT L (alignment length in basepairs: length of the aligned sequence)>=50

**Figure 2:**
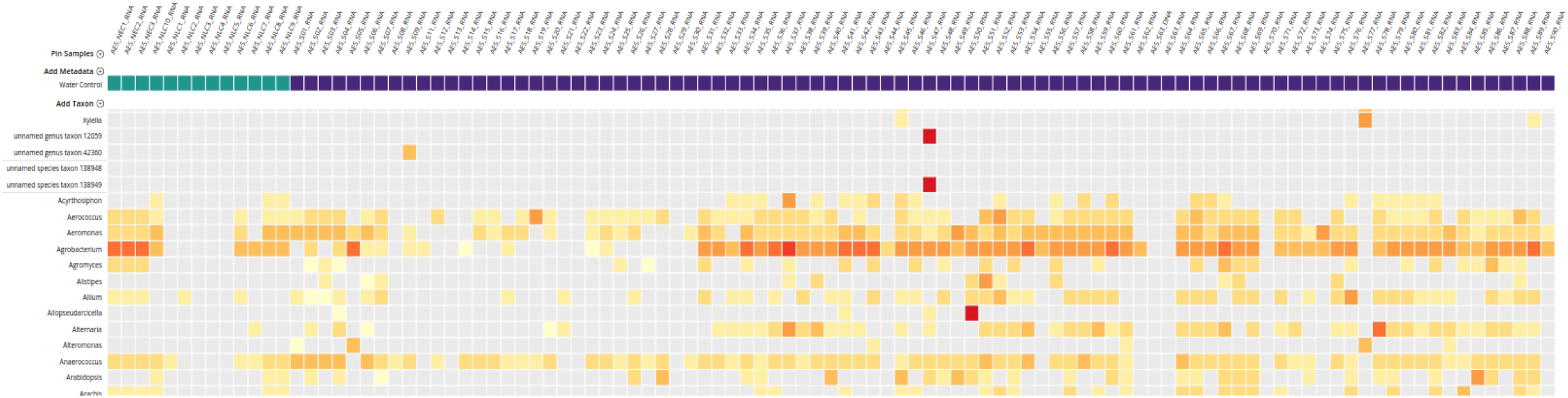
Heatmap depicting hits for Enterovirus (unnamed genus taxon 12059 and 138949) from sequencing of AES_S47_RNA. The names of the samples are on the top of the heat map. The samples marked by green are water controls during extraction (NEC) and library preparation (NLC). The heatmap was generated using the threshold of NT rPM (nucleotide reads per million) >=10 and NT L (alignment length in basepairs: length of the aligned sequence)>=50

Interestingly, only AES_S47_RNA showed a hit to Enterovirus B (strain Human coxsackievirus B1). This hit was particular to sample 47 and not seen in any negative controls. The metrics such as rPM of 359,409.1 (provides information of the abundance of a specific microbe within the sample), NT L (depicts the length of aligned sequence in base pair), Z score of 99 (shows the significance of any hit compared to the background), coverage visualization (assess breadth and depths of reads) and id of 85.8% signify that the organism hit is highly similar to the reference organism.^30^ The figure below shows the abundance of Enterovirus B in Sample 47 (NT rPM >=10 and NT L >=50). The coverage breadth of this hit was 98.7% with depth of 700.4x as seen in Figure 3.

**Figure 3:**
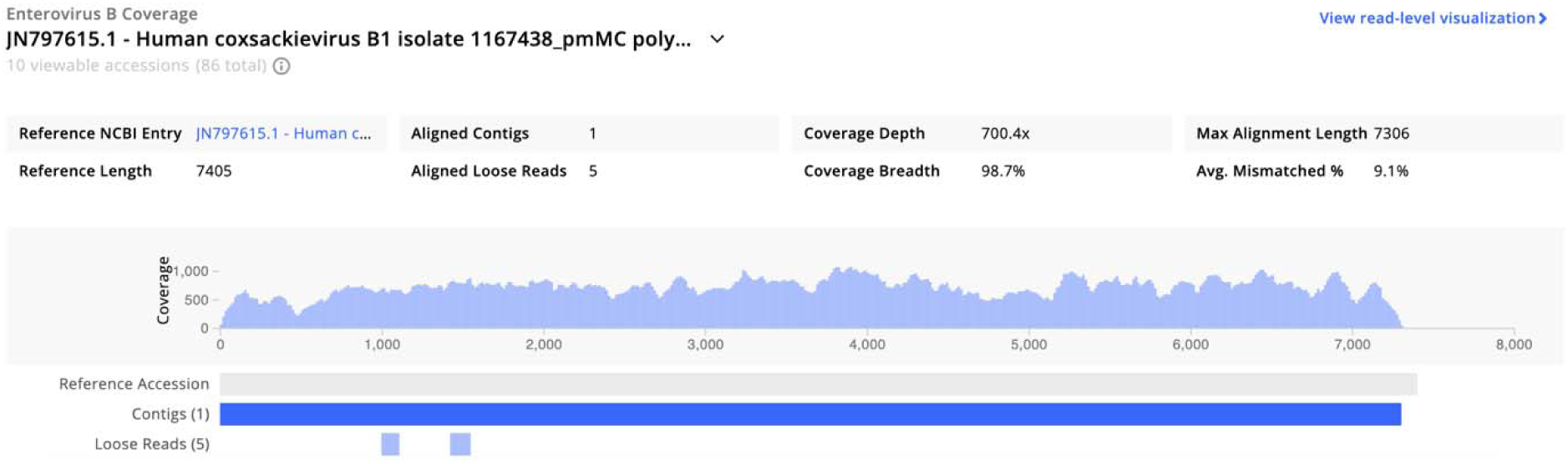
Coverage visualisation of Enterovirus in AES_S47_RNA from CZID, depicting the coverage metrics of the contigs generated for the particular hit along with coverage depth and width. The hit was visualised with the threshold of NT rPM >=10 and NT L >=50.

The strain Human coxsackievirus B1, from our study, was found similar to Coxsackievirus B1 responsible in mesangial renal disease.^28^ The genome similarly was also observed in genomes from coxsackievirus viruses causing myocarditis, severe gastroenteritis, food-and-mouth disease, respiratory distress, shown in Figure 4.^29–34^

**Figure 4:**
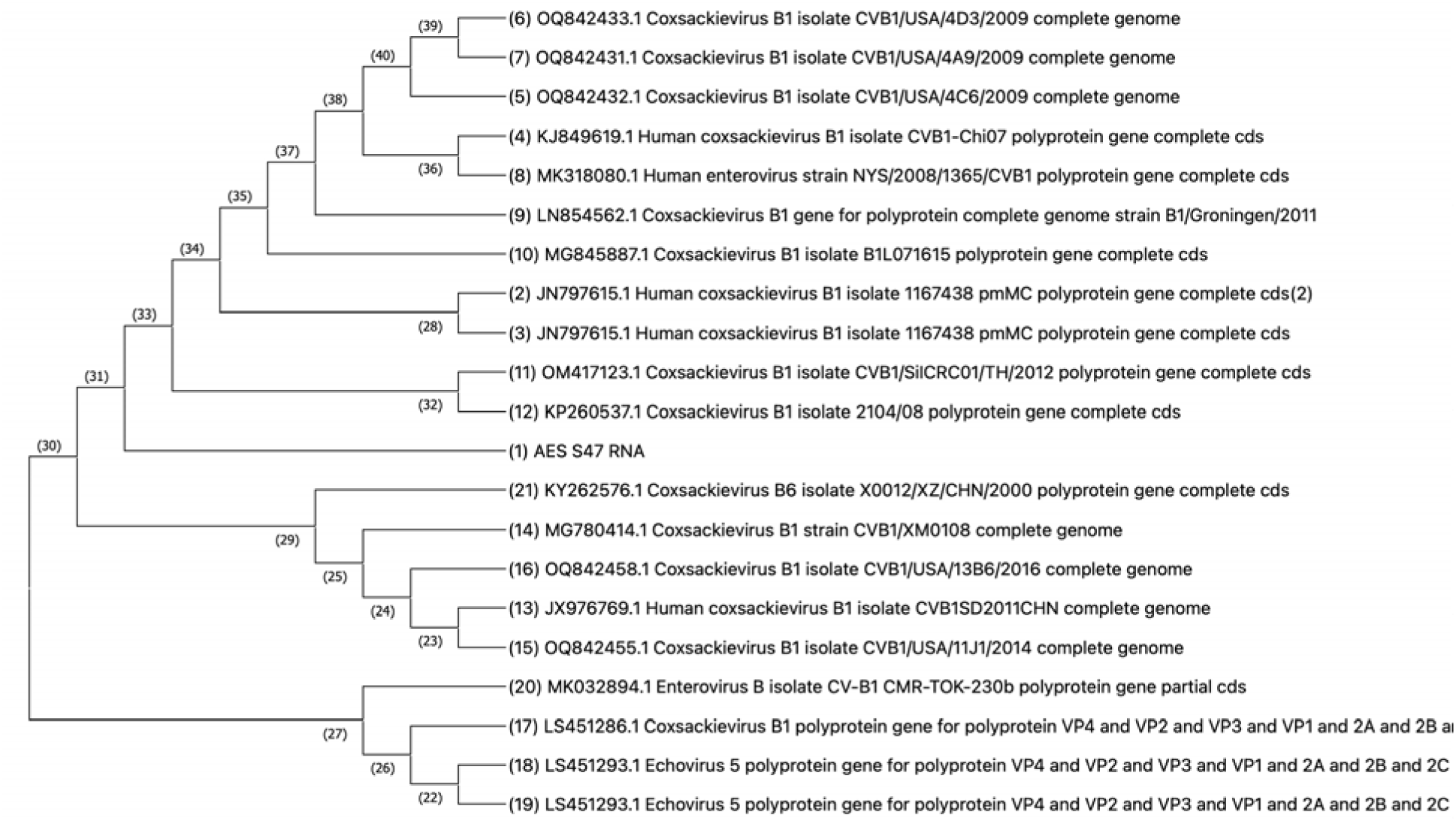
Phylogenetic comparison of AES_S47_RNA against other coxsackievirus B1 genomes, from NCBI. This tree was made using CLUSTAL W maximum likelihood statistical method, Tamura-Nei model with nearest neighbor interchange as the maximum likelihood heuristic method.

The strain Human coxsackievirus B1, from our study, was different (0.0569) from Enteroviruses B isolated from an outbreak in norther India, close in Nepal.^62^ (Figure 5)

**Figure 5:**
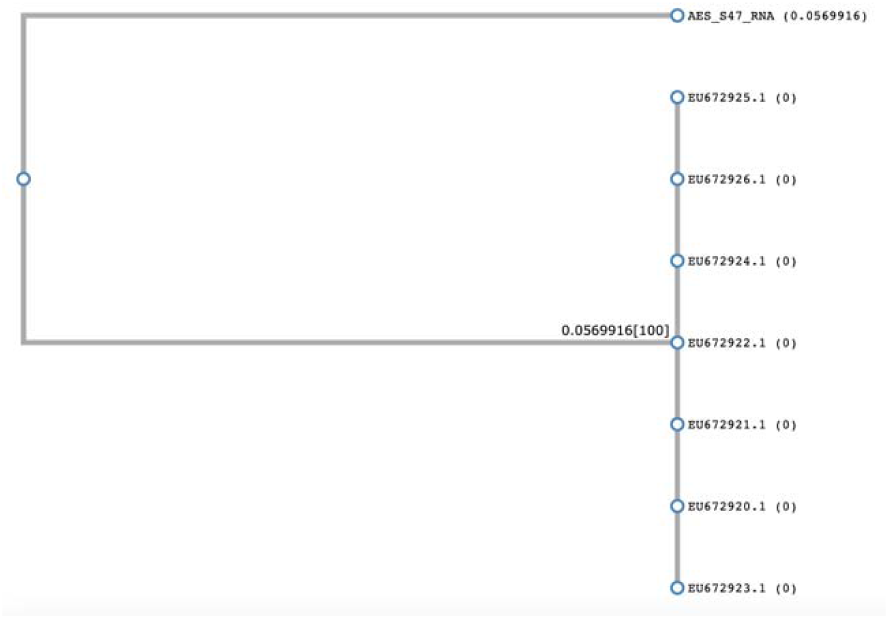
Phylogenic comparison of AES_S47_RNA against coxsackievirus B genomes isolated from an outbreak in I dia, based on based on partial 5’ noncoding region sequences. This tree was made using CLUSTALW.

**Figure 6:**
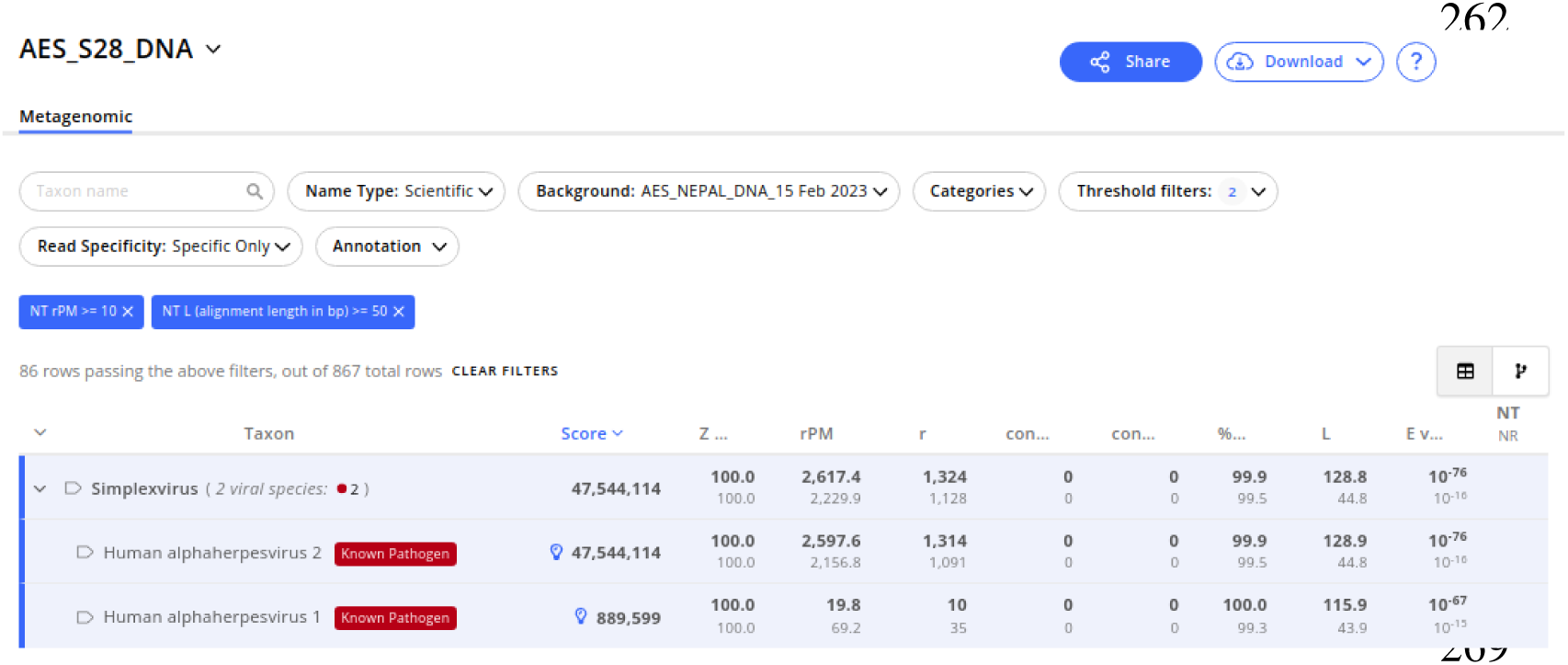
Result from CZID depicting the details of top hits in AES_S28_DNA along with various metrics related to the hit. The hits were visualised with the threshold of NT rPM >=10 and NT L >=50

### mNGS of DNA Libraries

In mNGS of DNA libraries, hits were observed for Human alphaherpesvirus 2 [AES_S28_DNA] from the first set. The same sample showed hit for Human alphaherpesvirus 1, but in a very low abundance, shown in figure 4. Additionally, background contaminants (laboratory and hospital) were seen in the water controls in this DNA sequencing result as well. Similar to RNA Libraries, most of the samples showed hits for Sphingomonas spp, Pseudomonas spp, and Acinetobacter spp. Nevertheless, the figure 5 depicts the result of the hit where there were 2,598.6 rPM for Human alphaherpesvirus 2.

However, due to lower coverage, contig visualization was not available for this hit.

### PCR Confirmation

#### Confirmation of Human alphaherpesvirus 2

Out of the two primer sets used, the established primers fared better providing Ct value of 23.11 for Human alphaherpesvirus 2.

#### Confirmation of Enterovirus B

After completion of snPCR for Enterovirus, the band was seen between 700-800bp after first amplification and between 300-400bp after final amplification. This confirmed the presence of Enterovirus as per the Enterovirus Surveillance Guidelines.^36^

## Discussion

### Demography of Acute Encephalitis Syndrome (AES)

Most of the subjects suffering from AES were young male population of median age 20 years. This gender distribution was concurrent to previous studies done, in Nepal, on epidemiology of AES.^37,38^ It has been observed that AES affects individuals from both gender and all ages, however, most of the studies have been done in younger population, as they pose high risk due to lack of developed antibodies.^39–42^ Another study done in Nepal also observed the young median age (19 years) for AES, while others observed older population.^37,38,43,44^

### Metagenomic Next Generation Sequencing

In this study, out of the 90 samples tested, most (n=88) of them could not be specified as specific hits. This was due to high level of background contaminants resulting in low confidence in calling organism hits within the experimental samples. Nevertheless, two samples showed confirmed hits for Enterovirus B and Human alphaherpesvirus 2, respectively, which differs from studies which depict that non-JE pathogens constitutes of 68-75% of AES cases.^7–10^ Nonetheless, the absence of causative agent in remaining samples could indicate that either the samples did not have intact nucleic to start with or had low pathogen abundance or could have been degraded because the ERCCs were amply sequenced from the RNA libraries.^16,45^

As per the result of mNGS, the high Z score (99) for Enterovirus B shows that hit for the organism is present significantly in our sample, when compared to the background. The average length of alignment (as shown by L metrics) is long (L=7258.7), which confirms for a good local alignment to reference.^46^ The id% is also higher (85.8%) meaning that the organism is highly similar to the reference organism in the database. Additionally, when the genome coverage is seen in detail, we can see that our sequenced genome depicts good coverage breadth and depth (depth of 700x and breadth of 98.7%), which is the range and uniformity of sequencing coverage for the particular hit.^45^ The presence of ENVB was also confirmed through snPCR followed by visualization of product size specific for all enteroviruses.^36^

The hit for alphaherpesvirus 2 was considered significant because it was not present in the control samples at the thresholds used to analyse the sample (high Z score of 100%, L value of 128.9, id% of 99.9%) considered reliable.^24,27,47^ The low contig value, for this hit, could be because of the organism being present at such a low abundance that the sequencer did not sequence enough reads to generate a contig. The contig value is dependent upon the total number of reads and the size of organism’s genome.^48^ Additionally, the decreased sensitivity of mNGS due to low abundance of pathogen has been studied for CSF.^49^ Several methods have been reported that can be used to increase the abundance of pathogen sequences or remove the unwanted host sequences.^50,51^ Nevertheless, as this genus is associated with encephalitis, the sample was taken further for analysis.^52,53^ During confirmation, the lower Ct value of 23.11 indicates presence of alphaherpesvirus 2, a known causative agent, in the sample.

### Enterovirus B and Human Alphaherpesvirus 2

Enterovirus B is a known causative agent of encephalitis.^16,54–56^ Enteroviruses are named by their transmission-route through the intestine.^57^ Studies have shown that enterovirus can cause various diseases in the nervous system, including aseptic meningitis, acute paralysis, encephalitis, meningo-encephalomyelitis among others, in children.^58–60^ Additionally, strain B1 has been documented to cause encephalomyocarditis (meningoencephalitis and severe myocarditis, often accompanied by heart failure) and showed genomic similarity to the enterovirus B from our study.^61^ Interestingly, studies in India have linked Enterovirus, among other pathogens, to AES, by various studies.^62–64^ For instance, Enterovirus outbreak was first reported from Uttar Pradesh, India in 2006 with seasonal outbreaks with high fatality occurring for several years.^62,65,66^ Southern Nepal borders with Uttar Pradesh, India and due to open borders and similar climate, it is plausible to find Enterovirus in CSF samples in Nepal. However, the strain of Enterovirus from our study was significantly different compared to genomes from the outbreak.^62^ Additionally, some studies in Nepal have reported Enterovirus as possible etiology of AES for Nepal.^67,68^

Similarly, Human alphaherpesvirus 2 is known to cause encephalitis in neonates and immunocompromised patients. Herpes simplex encephalitis (HSE) has significant morbidity and mortality, even with early diagnosis and treatment.^69,70^ HSV is found to be one of the predominant causes of AES in the western world.^71–73^ Among herpes simplex encephalitis, the vast majority of the encephalitis is caused by HSV-1, with HSV-2 being the etiology in less than 10% of the cases.^70^ Studies in India and Nepal have reported the presence of HSV-2 as causative agent of encephalitis, with varying range of incidence. ^69,74–78^

### Clinical Data and Process Control

However, due to lack of clinical metadata, the presence of Enterovirus B and Human alphaherpes 2 virus could not be clinically correlated. Clinical metadata such as onset of fever, date of infection, fatality, WBC counts, adjoining infection, etc are vital to correspond with the presence of infections.^16,79,80^

Additionally, usual environmental contaminants such as Sphingomonas spp., Pseudomonas spp, or Acinetobacter spp were seen. For instance, Sphingomonas are widely distributed in nature, having been isolated from many different land and water habitats, as well as from plant root systems, clinical specimens, and other sources. This is essentially due to their ability to survive in low concentrations of nutrients.^81,82^ Background contaminants of laboratory and hospital origin were also seen in the water controls. With appropriate use of background or negative controls, a background model can be created and subsequently subtracted from the results.^16,24^

### Collection Procedures

The lack of identification of causative agent in other 88 samples could be because all of samples that were analysed were as old as 2016 and 2017, and could possibly have gone through numerous freeze and thaw cycles. Therefore, the collection of samples in nucleic acid protectant such as Zymo RNA/DNA Shield would have protected the nucleic acid from degradation after sampling.^83,84^ Additionally, the causative agents could also have left the cerebrospinal fluid prior to collection depending upon the time of collection since the onset of fever, because it is advised to collect CSF within seven days of onset of fever.^85^

The possibility of freeze thaw cycles affecting the sample quality and lack of clinical metadata are limiting to the analysis, resulting in ambiguous interpretation of some samples. However, we contend that this aspect should not be corroborated as limitations, because the CSF samples analysed were not collected specifically for mNGS and there could be low abundance of the pathogen itself. Additionally, the sequencing was done in Illumina iSeq100 which has a maximum of approximately 4 million reads per run and can only accommodate a certain number of organisms with adequate coverage breadth and depth.^86^ Therefore, more deeper sequencing using sequencer with higher reads per run, host depletion and pathogen enrichment methods can be applied for samples with low pathogen abundance^.50,51^

## Conclusion

Identification and investigation of etiologies behind AES is essential for developing clinical management algorithms, improving surveillance with region-specific treatment and prevention policy as well as outbreak investigation. We do not expect the administration of mNGS as a regular diagnostic tool but rather an investigational and exploration tool to identify causative etiologies and develop molecular methods (such as qPCR) for diagnosis.

From this study, two documented, causative agents were revealed through metagenomic next generation sequencing and subsequently confirmed by PCR. Insufficiency of clinical metadata, process controls, and possibility of freeze thaw cycles affecting the sample quality incorporates ambiguity when correlating identified pathogens to infections. Therefore, there is a dire need of implementing standardized collection and storage procedures, including proper process controls and clinical metadata (WBC Count, primary diagnosis, discharge type, presence of another organism). Additionally, delicate samples such as CSF should be collected in a protectant and transported in a controlled and sterile environment.

## Data Availability

All data produced in the present study are available upon reasonable request to the author.
The pathogen genomic data can be found in Sequence Read Archive, National Center for Biotechnology Information (NCBI), under BioProject no PRJNA1019500.

## List of abbreviations

DNA: Deoxyribose Nucleic Acid
RNA: Ribo-Nucleic Acid
PCR: Polymerase Chain Reaction
AES: Acute Encephalitis Syndrome
WHO: World Health Organization
JEV: Japanese Encephalitis Virus
HSV: Herpes Simplex Virus
VSV: Varicella-Zoster Virus
mNGS: metagenomic Next Generation Sequencing
NGS: Next Generation Sequencing
LMIC: Low- and Middle-Income Countries
CSF: Cerebrospinal Fluid
WHO-IPD: World Health Organization Immunization Preventable Diseases
FWD: Family Welfare Department
NPHL: National Public Health Laboratory
NEB: New England Biolabs
ERCC: External Control Controls Consortium
CZ ID: Chan Zuckerberg ID
FP: Forward Primer
RP: Reverse Primer
snPCR: semi nested Polymerase Chain Reaction
NEC: Negative Extraction Control
NLC: Negative Library Control
NT: Nucleotide
rPM: Reads per Million
NT L: Nucleotide Length
Ct: Cycle of Threshold
HSE: Herpes Simplex Encephalitis
HSV: Herpes Simplex Virus

## Declarations

### Ethics approval and consent to participate

This study was ethically cleared from Nepal Health Research Council (NHRC) under id: 903 – 2019. This study directly did not contact the human subjects and investigated banked CSF samples and secondary metadata.

### Consent for publication

Not Applicable.

### Availability of data and materials

All data generated or analyzed during this study are included in this article. The pathogen genomic data can be found in Sequence Read Archive, National Center for Biotechnology Information (NCBI), under BioProject no PRJNA1019500. Further inquiries can be directed to the corresponding author.

### Authors’ contributions

Conceptualization, RS; methodology, RS and NK; investigation, RS and NK; resources, MV, CMT, VA, JG, NK; data curation, RS, NK, MV; writing-original draft preparation, RS and NK; writing-review and editing, DT, CMT, MV, VA, JG, SKM, BPG, RJ; supervision, RS; project administration, RS. All authors have read and agreed to the published version of the manuscript.

## Acknowledgements

We thank Bill and Melinda Gates Foundation, Grand Challenges Explorations Grant for the support. We appreciate the guidance from Chan Zuckerberg Biohub, San Francisco. We express our gratitude to Family Welfare Department and National Public Health Laboratory (NPHL) for supporting the study by providing the samples. We also thank WHO-IPD for providing data for the study.

## References

1. Guidelines for Surveillance of Acute Encephalitis Syndrome (With Special Reference to Japanese Encephalitis) NVBDCP. 2006. Accessed: 18 July 2022. Available at: http://www.nvbdcp.gov.in/Doc/AES%20guidelines.pdf

2. Granerod J, Crowcroft NS. The epidemiology of acute encephalitis. Neuropsychol Rehabil. 2007; 17:406–28.

3. Narain JP, Dhariwal AC, MacIntyre CR. Acute encephalitis in India: an unfolding tragedy. Indian J Med Res. 2017. 145:584–7.

4. Srivastava N, Deval H, Mittal M, Kant R, Bondre VP. The Outbreaks of Acute Encephalitis Syndrome in Uttar Pradesh, India (1978-2020) and Its Effective Management: A Remarkable Public Health Success Story. Front Public Health. 2022 Feb 9;9:793268.

5. Tripathy SK, Mishra P, Dwibedi B, Priyadarshini L, Das RR. Clinico-epidemiological Study of Viral Acute Encephalitis Syndrome Cases and Comparison to Nonviral Cases in Children from Eastern India. J Glob Infect Dis. 2019 Jan-Mar;11(1):7–12

6. National Health Mission. Routine Immunization, Government of India. Available at: http://www.nrhm.gov.in/nrhmcomponents/rmncha/immunization/background.html

7. AES/JE Cases and Deaths in the Country. National Vector Borne Disease Control Programme. Directorate General of Health Services. Ministry of Health and Family Welfare, Government of India. 2012.

8. Director, Child Health Division. Teku, Kathmandu, Nepal: Department of Health Services, Ministry of Health and Population. 2012. Acute Encephalitis Syndrome/Japanese Encephalitis Data of Nepal.

9. Cizman M, Jazbec J. Etiology of acute encephalitis in childhood in Slovenia. Pediatr Infect Dis J. 1993. 12:903–8.

10. Potharaju NR. Incidence Rate of Acute Encephalitis Syndrome without Specific Treatment in India and Nepal. Indian J Community Med. 2012 Oct. 37(4):240–51.

11. Joshi R, Kalantari SP, Reingold A et al. Changing landscape of acute encephalitis syndrome in India:A systematic review. Natl Med J India 2012. 25:212–20.

12. Rayamajhi A, Singh R, Prasad R, Khanal B, Singhi S: Study of Japanese encephalitis and other viral encephalitis in Nepali children. Pediatr Int. 2007, 49 (6): 978–984.

13. Pant GR: A serological survey of pigs, horses, and ducks in Nepal for evidence of infection with Japanese encephalitis virus. Ann N Y Acad Sci. 2006, 1081: 124–129.

14. Rayamajhi A, Ansari I, Ledger E, Bista KP, Impoinvil DE, Nightingale S, Kumar R, Mahaseth C, Solomon T, Griffiths MJ. Clinical and prognostic features among children with acute encephalitis syndrome in Nepal; a retrospective study. BMC Infect Dis. 2011 Oct 28;11:294

15. Hsu, V. P., Hossein, M. J., Parashar, U. D., Ali, M. M., Ksiazek, T. G., Kuzmin, I., Niezgoda, N., Rupprecht, C., Bresee, J., & Breiman, R. F. Nipah virus encephalitis reemergence, Bangladesh. Emerging Infectious Diseases. 2004. 10(12), 2082 – 2087.

16. Saha S, Ramesh A, Kalantar K, Malaker R, Hasanuzzaman M, Khan LM, Mayday MY, Sajib MSI, Li LM, Langelier C, Rahman H, Crawford ED, Tato CM, Islam M, Juan YF, de Bourcy C, Dimitrov B, Wang J, Tang J, Sheu J, Egger R, De Carvalho TR, Wilson MR, Saha SK, DeRisi JL. Unbiased Metagenomic Sequencing for Pediatric Meningitis in Bangladesh Reveals Neuroinvasive Chikungunya Virus Outbreak and Other Unrealized Pathogens. mBio. 2019 Dec 17;10(6):e02877–19.

17. Booss, J. & Esiri, M. M. Viral encephalitis in humans. 2003. Washington, DC: American Society for Microbiology Press.

18. Patricia J Simner, Steven Miller, Karen C Carroll, Understanding the Promises and Hurdles of Metagenomic Next-Generation Sequencing as a Diagnostic Tool for Infectious Diseases, Clinical Infectious Diseases, Volume 66, Issue 5, 1 March 2018, Pages 778–788

19. Ryan C. Shean & Alexander L. Greninger. One future of clinical metagenomic sequencing for infectious diseases, Expert Review of Molecular Diagnostics. 2019. 19:10, 849–851.

20. Phillips KA, Douglas MP, Marshall DA. Expanding Use of Clinical Genome Sequencing and the Need for More Data on Implementation. JAMA. 2020. Nov 24;324(20):2029–2030.

21. Gilbert GL. Molecular diagnostics in infectious diseases and public health microbiology: cottage industry to postgenomics. Trends Mol Med. 2002. 8(6):280–7.

22. Dwivedi, S., Purohit, P., Misra, R. et al. Diseases and Molecular Diagnostics: A Step Closer to Precision Medicine. Ind J Clin Biochem. 2007. 32, 374–398

23. Krishna NK, Cunnion KM. Role of molecular diagnostics in the management of infectious disease emergencies. Med Clin North Am. 2012. 96(6):1067–78.

24. Wilson MR, O’Donovan BD, Gelfand JM, Sample HA, Chow FC, Betjemann JP, Shah MP, Richie MB, Gorman MP, Hajj-Ali RA, Calabrese LH, Zorn KC, Chow ED, Greenlee JE, Blum JH, Green G, Khan LM, Banerji D, Langelier C, Bryson-Cahn C, Harrington W, Lingappa JR, Shanbhag NM, Green AJ, Brew BJ, Soldatos A, Strnad L, Doernberg SB, Jay CA, Douglas V, Josephson SA, DeRisi JL. Chronic meningitis investigated via metagenomic next-generation sequencing. JAMA Neurol. 2018. 75:947–955.

25. Langelier C, Kalantar KL, Moazed F, Wilson MR, Crawford E, Deiss T, Belzer A, Bolourchi S, Caldera S, Fung M, Jauregui A, Malcolm K, Lyden A, Khan L, Vessel K, Quan J, Zinter M, Chiu CY, Chow ED, Wilson J, Miller S, Matthay MA, Pollard KS, Christenson S, Calfee CS, DeRisi JL. Integrating host response and unbiased microbe detection for lower respiratory tract infection diagnosis in critically ill adults. Proc Natl Acad Sci USA. 2018. 115:E12353–E12362.

26. Wilson MR, Sample HA, Zorn KC, Arevalo S, Yu G, Neuhaus J, Federman S, Stryke D, Briggs B, Langelier C, Berger A, Douglas V, Josephson SA, Chow FC, Fulton BD, DeRisi JL, Gelfand JM, Naccache SN, Bender J, Dien Bard J, Murkey J, Carlson M, Vespa PM, Vijayan T, Allyn PR, Campeau S, Humphries RM, Klausner JD, Ganzon CD, Memar F, Ocampo NA, Zimmermann LL, Cohen SH, Polage CR, DeBiasi RL, Haller B, Dallas R, Maron G, Hayden R, Messacar K, Dominguez SR, Miller S, Chiu CY. Clinical Metagenomic Sequencing for Diagnosis of Meningitis and Encephalitis. N Engl J Med. 2019 Jun 13;380(24):2327–2340.

27. Kalantar KL, Carvalho T, de Bourcy CFA, Dimitrov B, Dingle G, Egger R, Han J, Holmes OB, Juan YF, King R, Kislyuk A, Lin MF, Mariano M, Morse T, Reynoso LV, Cruz DR, Sheu J, Tang J, Wang J, Zhang MA, Zhong E, Ahyong V, Lay S, Chea S, Bohl JA, Manning JE, Tato CM, DeRisi JL. IDseq-An open source cloud-based pipeline and analysis service for metagenomic pathogen detection and monitoring. Gigascience. 2020 Oct 15;9(10):giaa111.

28. Namvar L, Olofsson S, Bergström T, Lindh M. Detection and typing of Herpes Simplex virus (HSV) in mucocutaneous samples by TaqMan PCR targeting a gB segment homologous for HSV types 1 and 2. J Clin Microbiol. 2005 May;43(5):2058–64.

29. Bachtler, M., Frey, B.M., Frey, F.J., Gorgievski, M., Simonetti, G. and Pasch, A Role of enteroviruses in mesangial renal disease [Unpublished]

30. Quinn KK, Wollersheim SK, Krogstad P. Complete Genome Sequence of Coxsackievirus B1 Isolated during Case Outbreaks in 2007 in the United States. Genome Announc. 2014 Jul 24;2(4):e00574–14. doi: 10.1128/genomeA.00574-14. PMID: 25059857; PMCID: PMC4110215.

31. Bavelaar HH, Rahamat-Langendoen J, Niesters HG, Zoll J, Melchers WJ. Whole genome sequencing of fecal samples as a tool for the diagnosis and genetic characterization of norovirus. J Clin Virol. 2015 Nov;72:122–5. doi: 10.1016/j.jcv.2015.10.003. Epub 2015 Nov 6. PMID: 26492615.]

32. Prasertsopon, J., Sangsiriwut, K., Noisumdaeng, P., Buathong, R. and Puthavathana, P. Enterovirus-associated hand, foot and mouth diseases in Thailand JOURNAL Unpublished. Submitted (26-JAN-2022)]

33. Chehadeh, W., Maimoona, S. and Kurien, S.S. Molecular characterization of coxsackievirus B1 isolated from 1-year-old child with respiratory distress. JOURNAL Unpublished. Submitted 7 Dec 2014.

34. Zhang T, Du J, Xue Y, Su H, Yang F, Jin Q. Epidemics and Frequent Recombination within Species in Outbreaks of Human Enterovirus B-Associated Hand, Foot and Mouth Disease in Shandong China in 2010 and 2011. PLoS One. 2013 Jun 19;8(6):e67157. doi: 10.1371/journal.pone.0067157. PMID: 23840610; PMCID: PMC3686723.

35. CZID Portal. https://czid.org. Accessed July 2022

36. WHO. Enterovirus Surveillance Guidelines. WHO Regional Office for Europe. 2015. Available at: https://www.euro.who.int/data/assets/pdf_file/0020/272810/EnterovirusSurveillanceGuidelines.pdf

37. Rayamajhi, et al. Evaluating cognitive outcomes in adult patients with acute encephalitis syndrome: a prospective study from a tertiary care center in Nepal. Encephalitis. 2022; 2(2): 36–44

38. Thapa, et al. Clinical profile and outcome of acute encephalitis syndrome (AES) patients treated in College of Medical Sciences-Teaching Hospital. Journal of College of Medical Sciences. 2013. Vol 9, No.2, 31–37.)

39. Rayamajhi, A., Ansari, I., Ledger, E. et al. Clinical and prognostic features among children with acute encephalitis syndrome in Nepal; a retrospective study. BMC Infect Dis 11, 294 (2011). 10.1186/1471-2334-11-294 AND

40. Griffiths MJ, Lemon JV, Rayamajhi A, Poudel P, Shrestha P, Srivastav V, et al. (2013) The Functional, Social and Economic Impact of Acute Encephalitis Syndrome in Nepal – a Longitudinal Follow-Up Study. PLoS Negl Trop Dis 7(9): e2383. AND

41. Roy DB, Khatri HV. Study of Demographic Profile, Etiology, and Clinical Outcome in Patients Admitted With Acute Encephalitis Syndrome From the Western Part of India. Cureus. 2022 Mar 11;14(3):e23085. doi: 10.7759/cureus.23085. PMID: 35464588; PMCID: PMC9001832. And

42. Jmor, F., Emsley, H.C., Fischer, M. et al. The incidence of acute encephalitis syndrome in Western industrialised and tropical countries. Virol J 5, 134 (2008). 10.1186/1743-422X-5-134)

43. Clinical presentation, etiology, and survival in adult acute encephalitis syndrome in rural Central India. Joshi R, Mishra PK, Joshi D, et al. 10.1016/j.clineuro.2013.04.008. Clin Neurol Neurosurg. 2013;115:1753–1761.

44. Roy DB, Khatri HV. Study of Demographic Profile, Etiology, and Clinical Outcome in Patients Admitted With Acute Encephalitis Syndrome From the Western Part of India. Cureus. 2022 Mar 11;14(3):e23085. doi: 10.7759/cureus.23085. PMID: 35464588; PMCID: PMC9001832.

45. Thomas T, Gilbert J, Meyer F. Metagenomics - a guide from sampling to data analysis. Microb Inform Exp. 2012 Feb 9;2(1):3.

46. CZID Portal. h https://chanzuckerberg.zendesk.com/hc/en-us. Accessed July 2022.

47. Zinter MS, Dvorak CC, Mayday MY, Iwanaga K, Ly NP, McGarry ME, Church GD, Faricy LE, Rowan CM, Hume JR, Steiner ME, Crawford ED, Langelier C, Kalantar K, Chow ED, Miller S, Shimano K, Melton A, Yanik GA, Sapru A, DeRisi JL. Pulmonary Metagenomic Sequencing Suggests Missed Infections in Immunocompromised Children. Clin Infect Dis. 2019 May 17;68(11):1847–1855

48. Ayling M, Clark MD, Leggett RM. New approaches for metagenome assembly with short reads. Brief Bioinform. 2020 Mar 23;21(2):584–594.

49. Miller S, Naccache SN, Samayoa E, Messacar K, Arevalo S, Federman S, Stryke D, Pham E, Fung B, Bolosky WJ, Ingebrigtsen D, Lorizio W, Paff SM, Leake JA, Pesano R, DeBiasi R, Dominguez S, Chiu CY. Laboratory validation of a clinical metagenomic sequencing assay for pathogen detection in cerebrospinal fluid. Genome Res. 2019. 29:831–842.

50. Gu W, Crawford ED, O’Donovan BD, Wilson MR, Chow ED, Retallack H, DeRisi JL. Depletion of abundant sequences by hybridization (DASH): using Cas9 to remove unwanted high-abundance species in sequencing libraries and molecular counting applications. Genome Biol. 2016. 17:41.

51. Quan J, Langelier C, Kuchta A, Batson J, Teyssier N, Lyden A, Caldera S, McGeever A, Dimitrov B, King R, Wilheim J, Murphy M, Ares LP, Travisano KA, Sit R, Amato R, Mumbengegwi DR, Smith JL, Bennett A, Gosling R, Mourani PM, Calfee CS, Neff NF, Chow ED, Kim PS, Greenhouse B, DeRisi JL, Crawford ED. FLASH: a next-generation CRISPR diagnostic for multiplexed detection of antimicrobial resistance sequences. Nucleic Acids Res. 2019. 47:e83.

52. Carneiro, V.C.d., Alves-Leon, S.V., Sarmento, D.J.d., et al. Herpesvirus and neurological manifestations in patients with severe coronavirus disease. Virol J 2022. 19, 10.

53. Patil S, Beck P, Nelson TB, Bran A, Roland W. Herpes Simplex Virus-2 Meningoencephalitis With Abducens Nerve Palsy With Literature Review. Cureus. 2021 Jun 8;13(6):e15523

54. Kumar A, Shukla D, Kumar R, Idris MZ, Misra UK, Dhole TN. Molecular epidemiological study of enteroviruses associated with encephalitis in children from India. J Clin Microbiol. 2012. 50:3509– 3512.

55. Jain S, Patel B, Bhatt GC. Enteroviral encephalitis in children: clinical features, pathophysiology, and treatment advances. Pathog Glob Health. 2014 Jul;108(5):216–22

56. Calvo C, Gallardo P, Torija P, Bellón S, Méndez-Echeverría A, Del Rosal T, Baquero-Artigao F, Sainz T, Romero M, Cabrerizo M. Enterovirus neurological disease and bacterial coinfection in very young infants with fever. J Clin Virol. 2016 Dec;85:37–39

57. “Genus: Enterovirus”. International Committee on Taxonomy of Viruses (ICTV). Accessed: 18 July 2022. Derivation of names Entero: from Greek enteron, ‘intestine’

58. Chaudhary MC, Bronze MS. Enterovirus Infection. Medscape: Infectious Diseases. 2019. Accessed: 18 July 2022.

59. Glaser CA, Gilliam S, Schnurr D et al. In search of encephalitis etiologies: diagnostic challenges in the California Encephalitis Project, 1998–2000. Clin. Infect. Dis. 2003. 36(6), 731–742.

60. Gofshteyn J, Cárdenas AM, Bearden D. Treatment of chronic enterovirus encephalitis with fluoxetine in a patient with X-linked agammaglobulinemia. Pediatr. Neurol. 2016. 64, 94–98.

61. Wikswo ME, Khetsuriani N, Fowlkes AL, Zheng X, Peñaranda S, Verma N, Shulman ST, Sircar K, Robinson CC, Schmidt T, Schnurr D, Oberste MS. Increased activity of Coxsackievirus B1 strains associated with severe disease among young infants in the United States, 2007-2008. Clin Infect Dis. 2009 Sep 1;49(5):e44–51. doi: 10.1086/605090. PMID: 19622041

62. Sapkal GN, Sapkal GN, Bondre VP, Fulmali PV. Enteroviruses in patients with acute encephalitis, Uttar pradesh, India. Emerg Infect Dis. 2009. 15: 295–298.

63. Mittal M, Kushwaha KP, Pandey AK, Fore MM. A clinico-epidemiological study of acute encephalitis syndrome with multi organ dysfunction. International Journal of Contemporary Pediatrics. 2017. 4(3).

64. Ravi V, Hameed SKS, Desai A, Mani RS, Reddy V, Velayudhan A, Yadav R, Jain A, Saikia L, Borthakur AK, Sharma A, Mohan DG, Bhandopadhyay B, Bhattacharya N, Inamdar L, Hossain S, Daves S, Sejvar J, Dhariwal AC, Sen PK, Venkatesh S, Prasad J, Laserson K, Srikantiah P. An algorithmic approach to identifying the aetiology of acute encephalitis syndrome in India: results of a 4-year enhanced surveillance study. Lancet Glob Health. 2022 May;10(5):e685–e693.

65. Mittal M, Bondre V, Murhekar M, Deval H, Rose W, Verghese VP, Mittal M, Patil G, Sabarinathan R, Vivian Thangaraj JW, Kanagasabai K, Prakash JAJ, Gupta N, Gupte MM, Gupte MD. Acute Encephalitis Syndrome in Gorakhpur, Uttar Pradesh, 2016: Clinical and Laboratory Findings. Pediatr Infect Dis J. 2018 Nov;37(11):1101–1106

66. Kumar A, Shukla D, Kumar R, Idris MZ, Misra UK, Dhole TN. An epidemic of encephalitis associated with human enterovirus B in Uttar Pradesh, India, 2008. J Clin Virol. 2011 Jun;51(2):142–5.

67. Giri A, Arjyal A, Koirala S, Karkey A, Dongol S, Thapa SD, Shilpakar O, Shrestha R, van Tan L, Thi Thuy Chinh BN, Krishna K C R, Pathak KR, Shakya M, Farrar J, Van Doorn HR, Basnyat B. Aetiologies of central nervous system infections in adults in Kathmandu, Nepal: a prospective hospital-based study. Sci Rep. 2013;3:2382

68. Säll O, Thulin Hedberg S, Neander M, Tiwari S, Dornon L, Bom R, Lagerqvist N, Sundqvist M, Mölling P. Etiology of Central Nervous System Infections in a Rural Area of Nepal Using Molecular Approaches. Am J Trop Med Hyg. 2019 Jul;101(1):253–259.

69. Bookstaver PB, Mohorn PL, Shah A, Tesh LD, Quidley AM, Kothari R, Bland CM, Weissman S. Management of viral central nervous system infections: a primer for clinicians. J Cent Nerv Syst Dis. 2017. 9.

70. AK AK, Mendez MD. Herpes Simplex Encephalitis. 2022 Mar 15. In: StatPearls [Internet]. Treasure Island (FL): StatPearls Publishing; 2022 Jan–. PMID: 32491575.

71. Khetsuriani, N., Holman, R. C. & Anderson, L. J. Burden of encephalitis-associated hospitalizations in the United States, 1988–1997. Clinical infectious diseases: an official publication of the Infectious Diseases Society of America. 2002. 35. 175–182.

72. Davison, K. L., Crowcroft, N. S., Ramsay, M. E., Brown, D. W. & Andrews, N. J. Viral encephalitis in England, 1989–1998: what did we miss? Emerging infectious diseases. 2003. 9. 234–240.

73. Huppatz, C. et al. Etiology of encephalitis in Australia, 1990–2007. Emerging infectious diseases. 2009. 15. 1359–1365.

74. Panagariya A, Jain RS, Gupta S, Garg A, Sureka RK, Mathur V. Herpes simplex encephalitis in North West India. Neurol India. 2001 Dec;49(4):360–5

75. Bradshaw MJ, Venkatesan A. Herpes Simplex Virus-1 Encephalitis in Adults: Pathophysiology, Diagnosis, and Management. Neurotherapeutics. 2016 Jul;13(3):493–508.

76. Kubo T, Rai SK, Nakanishi M, Yamano T. Seroepidemiological study of herpes viruses in Nepal. Southeast Asian J Trop Med Public Health. 1991 Sep;22(3):323–5

77. Adhikari A. A Case Report on Herpes Simplex Encephalitis. Medical Journal of Shree Birendra Hospital. 2012. 11(1).

78. Rayamajhi P, Nepal G, Ojha R, Rajbhandari R, Gajurel BP, Karn R. Evaluating cognitive outcomes in adult patients with acute encephalitis syndrome: a prospective study from a tertiary care center in Nepal. 2022. 2(2). 36–44.

79. Ramesh A, Nakielny S, Hsu J, Kyohere M, Byaruhanga O, de Bourcy C, Egger R, Dimitrov B, Juan YF, Sheu J, Wang J, Kalantar K, Langelier C, Ruel T, Mpimbaza A, Wilson MR, Rosenthal PJ, DeRisi JL. Metagenomic next-generation sequencing of samples from pediatric febrile illness in Tororo, Uganda. PLoS One. 2019 Jun 20;14(6):e0218318.

80. Institute of Medicine (US) Roundtable on Value & Science-Driven Health Care. Clinical Data as the Basic Staple of Health Learning: Creating and Protecting a Public Good: Workshop Summary. Washington (DC): National Academies Press (US); 2010. Available from: https://www.ncbi.nlm.nih.gov/books/NBK54302/

81. Leys NM, Ryngaert A, Bastiaens L, Verstraete W, Top EM, Springael D. Occurrence and phylogenetic diversity of Sphingomonas strains in soils contaminated with polycyclic aromatic hydrocarbons. Appl Environ Microbiol. 2004 Apr;70(4):1944–55

82. Cavicchioli R, Fegatella F, Ostrowski M, Eguchi M, Gottschal J. Sphingomonads from marine environments. J Ind Microbiol Biotechnol. 1999 Oct;23(4-5):268–272.

83. Thraenhart O, Jursch C. Virucidal activity if the nucleic acid preservation product “DNA/RNA Shield” against the murine parvovirus (MVM) at 20°C. 2018. Eurovir.

84. Phommanivong, V., Kanda, S., Shimono, T. et al. Co-circulation of the dengue with chikungunya virus during the 2013 outbreak in the southern part of Lao PDR. Trop Med Health. 2016. 44. 24

85. New York State Department of Health. Collection and Submission of Specimens for Viral Encephalitis Testing. July 2010.

86. seq100 Specifications. Accessed: 18 July 2022. Available at https://support.illumina.com/bulletins/2020/04/maximum-read-length-for-illumina-sequencing-platforms.html

